# Serological surveys to estimate cumulative incidence of SARS-CoV-2 infection in adults (Sero-MAss study), Massachusetts, July-August 2020: a mail-based cross-sectional study

**DOI:** 10.1101/2021.03.05.21249174

**Authors:** Teah Snyder, Johanna Ravenhurst, Estee Y. Cramer, Nicholas G. Reich, Laura B. Balzer, Dominique Alfandari, Andrew A. Lover

## Abstract

**Background:** The SARS-CoV-2 pandemic is an unprecedented global health crisis. The state of Massachusetts was especially impacted during the initial stages; however, the extent of asymptomatic transmission remains poorly understood due to limited asymptomatic testing in the “first wave.” To address this gap, a geographically representative and contact-free seroprevalence survey was conducted in July-August 2020, to estimate prior undetected SARS-CoV-2 infections.

**Methods:** Students, faculty, librarians and staff members at the University of Massachusetts, Amherst without a previous COVID-19 diagnosis were invited to participate in this study along with one member of their household in June 2020. Two separate sampling frames were generated from administrative lists: all undergraduates and their household members (primary sampling group) were randomly selected with probability proportional to population size. All staff, faculty, graduate students and librarians (secondary sampling group) were selected as a simple random sample. After informed consent and a socio-behavioral survey, participants were mailed test kits and asked to return self-collected dried blood spot (DBS) samples. Samples were analyzed via ELISA for anti-SARS-CoV-2 IgG antibodies, and then IgM antibodies if IgG-positive. Seroprevalence estimates were adjusted for survey non-response. Binomial models were used to assess factors associated with seropositivity in both sample groups separately.

**Results:** Approximately 27,000 persons were invited via email to assess eligibility. Of the 1,001 individuals invited to participate in the study, 762 (76%) returned blood samples for analysis. In the primary sampling group 548 returned samples, of which 230 enrolled a household member. Within the secondary sampling group of 214 individuals, 79 enrolled a household member. In the primary sample group, 36 (4.6%) had IgG antibodies detected for an estimated weighed prevalence for this population of 5.3% (95% CI: 3.5 to 8.0). In the secondary sampling group, 10 (3.4%) of 292 individuals had IgG antibodies detected for an estimated adjusted prevalence of 4.0% (95% CI: 2.2 to 7.4). No samples were IgM positive. No association was found in either sample group between seropositivity and self-reported work duties or customer-facing hours. In the primary sampling group, self-reported febrile illness since Feb 2020, male sex, and minority race (Black or American Indian/Alaskan Native) were associated with seropositivity. No factors except geographic regions within the state were associated with evidence of prior SARS-CoV-2 infection in the secondary sampling group.

**Interpretation:** This study provides insight into the seroprevalence of university-related populations and their household members across the state of Massachusetts during the summer of 2020 of the pandemic and helps to fill a critical gap in estimating the levels of sub-clinical and asymptomatic infection. Estimates like these can be used to calibrate models that estimate levels of population immunity over time to inform public health interventions and policy.

**Funding:** UMass Faculty Fund (A Lover); UMass Faculty Discretionary Funds (N Reich); UMass Institute for Applied Life Science “Midigrant” (#169076; A. Lover); and D. Alfandari was supported by grants from the USPHS (R24OD021485).

## Introduction

Since emergence in early 2020, the SARS-CoV-2 virus has severely impacted the entire globe. The state of Massachusetts was heavily impacted in the earliest stages of the pandemic, and a “super-spreader” event in the state in April 2020 may have seeded large case clusters throughout the country.^1^ However, the trajectory of the early stages of transmission in this state, as well as across the US remain poorly understood due to changes in case definitions and limited testing of both symptomatic and asymptomatic persons during the summer of 2020.^2^ To assess seroprevalence across the state, a mail-based serosurvey was implemented July-August 2020. At the time of this survey, the Massachusetts Department of Public Health had reported over 109,143 confirmed COVID-19 cases and over 8,081 deaths.^3^ Seroepidemiological studies are a critical tool to explore infection dynamics, especially where asymptomatic or subclinical infections are common, as for SARS-CoV-2.^4^ This study helps to fill a critical gap in estimating the levels of sub-clinical and asymptomatic infection to inform consequent levels of population-immunity.^5^

Concurrent to this study, a number of seroprevalence studies were conducted on the east coast of the United States; these studies focused on specific populations at high risk and found varying results. A survey in April 2020 in a convenience sample of 200 asymptomatic residents of Chelsea, MA found an estimated of seroprevalence of 31.5% (17.5% IgM+/IgG+, 9.0% IgM+/IgG- and 5.0% IgM-/IgG+).^6^ This study used a small convenience sample and did not include any randomization.^6^ A study with a larger sample of over 28,000 clinical patient samples in New York City, USA found an IgG seropositivity prevalence of 44% with over 50% of participants reporting no symptoms.^7^

Other seroprevalence surveys across the US have found generally low- to moderate prevalence in a diverse set of study populations. A study of 790 university students in Los Angeles, California conducted in April and May of 2020 estimated a prevalence of SARS-CoV-2 IgG antibody of 4.0% (95% CI: 3.0 to 5.1%).^8^ During May – April of 2020, a cross-sectional study in St. Louis found IgG seropositivity to be estimated at 1.71% (95% CI 0.04% to 3.38%) in pediatric patients and 3.11% (95% CI: 0.92% to 5.32%) in adult patients. In the most comprehensive serosurvey from the spring and summer months of 2020, 16,025 clinical samples were analyzed with IgG spike protein sero-reactivity ranging from 1.0% in the San Francisco Bay Area to 6.9% in New York City.^9^ These disparate results highlight major geographic variability in the trajectory of infections, and reinforce the need for additional seroprevalence studies to more fully contextualize trends in immunity to SARS-CoV-2 targeting specific geographic regions.

Though community seroprevalence studies generally rely on serum samples collected in health facilities, the use of dried blood spot (DBS) samples is a practical and effective alternative.^10^ DBS samples involve a small finger-prick sample self-collected by participants in their own homes. The use of dried blood samples for antibody assays has been validated in other work prior to the current pandemic,^10,11^ and previous studies have evaluated the feasibility, validity, and acceptability of using DBS samples for SARS-CoV-2 antibody testing.^12–15^ This method of sample collection facilitates efficient population-level sampling while minimizing social mixing and concurrent potential exposures.

This study estimated the prevalence of previous infection with SARS-CoV-2 in individuals who had not been diagnosed with COVID-19 and were asymptomatic with representative coverage across the entire state of Massachusetts, USA. Information from this study can provide knowledge regarding the seropositivity of this population and can be used to inform decision-making regarding community re-openings during the pandemic.

## Methods

### Study Design and Participants

The study population included undergraduate students, graduate students, staff, and faculty members currently affiliated with the University of Massachusetts, Amherst and their household members. On-campus classes were suspended in mid-March 2020; consequently, undergraduates had exposure to the local epidemiology within their communities from March until sampling in July-August throughout the state (primary sampling group). Conversely, graduate students, faculty, staff, librarians and their family members (secondary sampling group) almost universally reside in close proximity to Amherst, and broadly reflect transmission in the Western part of the state. UMass affiliates were eligible to participate in this study if they were above the age of 18, had been living in Massachusetts for the past eight weeks; had never received a COVID-19 diagnosis from a medical professional; and did not have a fever greater than 100.4° F at the time of survey completion. Household members were eligible for inclusion if they met all of these same criteria and were between the ages of 23 and 78 (chosen to expand sampling beyond college-age population groups).

An institutional email list was provided by university administration for recruitment. Initial emails were sent out to UMass affiliates between June 23, 2020 and June 26, 2020 for participant recruitment. The email provided information about the study and links to a screening eligibility survey, informed consent document, initial survey regarding COVID-19 risk factors, and information regarding shipping addresses. If the UMass affiliate had a household member interested in participating, a single household member was invited to complete an eligibility, consent, and initial survey forms. To increase participation rates, two reminder emails were sent to all non-respondents (day three and six after initial solicitation). All survey responses were collected and stored in REDCap.^16^

The survey was closed after a three-week enrollment period, and a subset of participants were selected to receive a test kit. To select a population representative of the broader UMass community across the entire state of Massachusetts, two sampling schemes were applied. The first consisted of all undergraduates and their household members (<u>primary sampling group</u>); the second sampling frame consisted of graduate students, staff, faculty members, librarians, and their household members (<u>secondary sampling group</u>). Within the primary sampling group, selection for biosample collection used probability proportional to population size, using the most recent census data aggregated to state-level emergency response regions due to sparse county-level populations (Figure 1).^17^ For the secondary sampling group, selection for biosampling was via simple random sampling.

**Fig 1.**
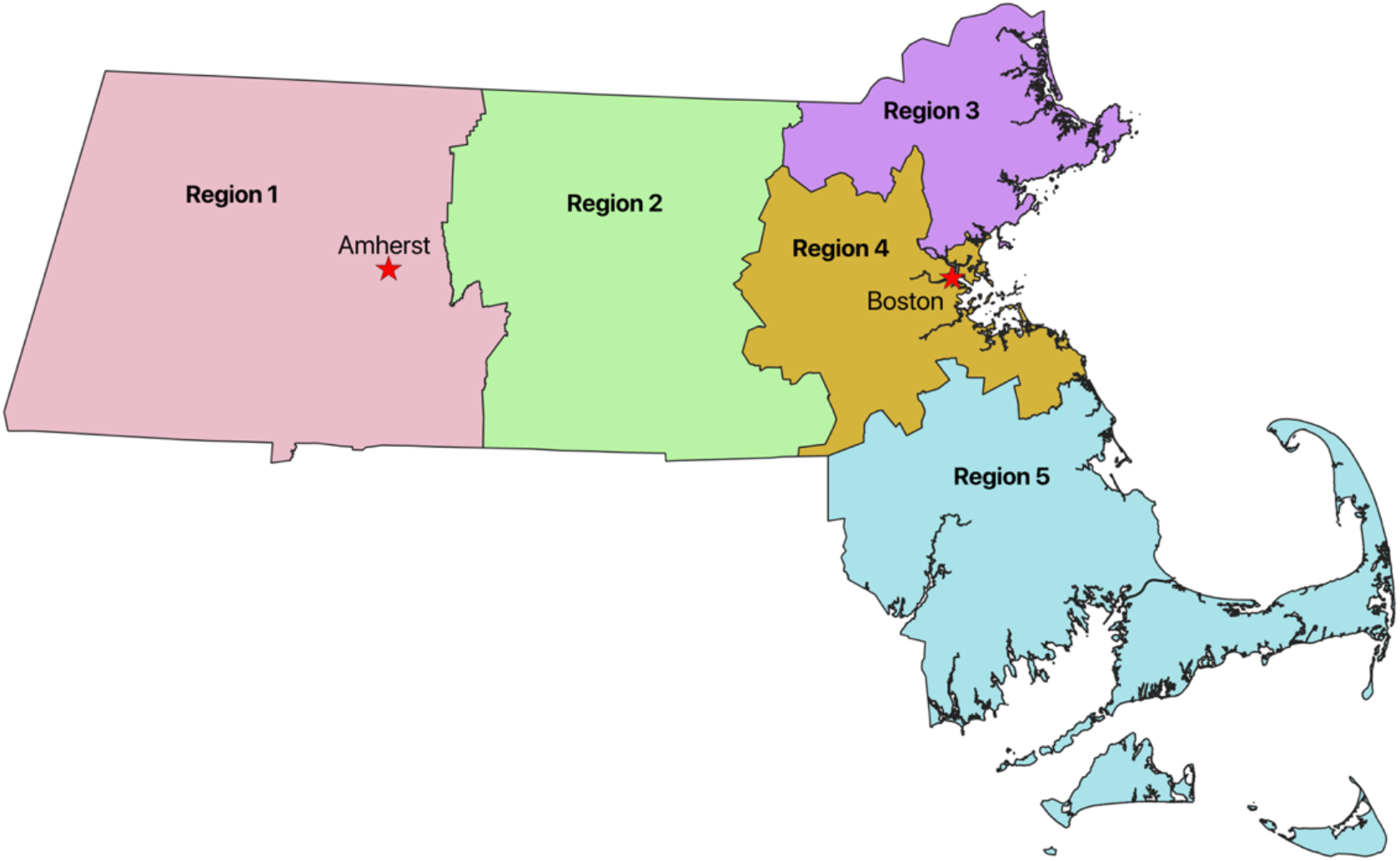
Emergency Response Region sampling frames, for SARS-CoV-2 seropositivity, Massachusetts, USA, Jul-Aug 2020.

The full sample frame selection is shown in Figure 2. Briefly, an email invitation was sent to a total of 27,339 individuals, of which 4,124 completed the screening, informed consent, and initial survey forms. A total of 1,001 individuals were then randomly selected to receive a sampling kit. Participants were mailed all materials to safely collect and return samples, including lancets, alcohol wipes, gauze, gloves, bandages, a bloodspot collection card, a pre-paid shipping box, and detailed printed instructions cards (including a nurse call line). Upon mailing out the test kits, participants were also emailed a link containing a video on how to collect the DBS, along with a detailed survey form with demographics, risk factors, and any current symptoms or COVID diagnoses. No participants reported a COVID-19 diagnosis between the initial survey, and sample collection several weeks later. All shipments utilized a Biological Substance Category B (UN3373) shipping box. This study was approved by the UMass Amherst Human Research Protection Office (Approval #2062; April 27, 2020).

**Fig 2.**
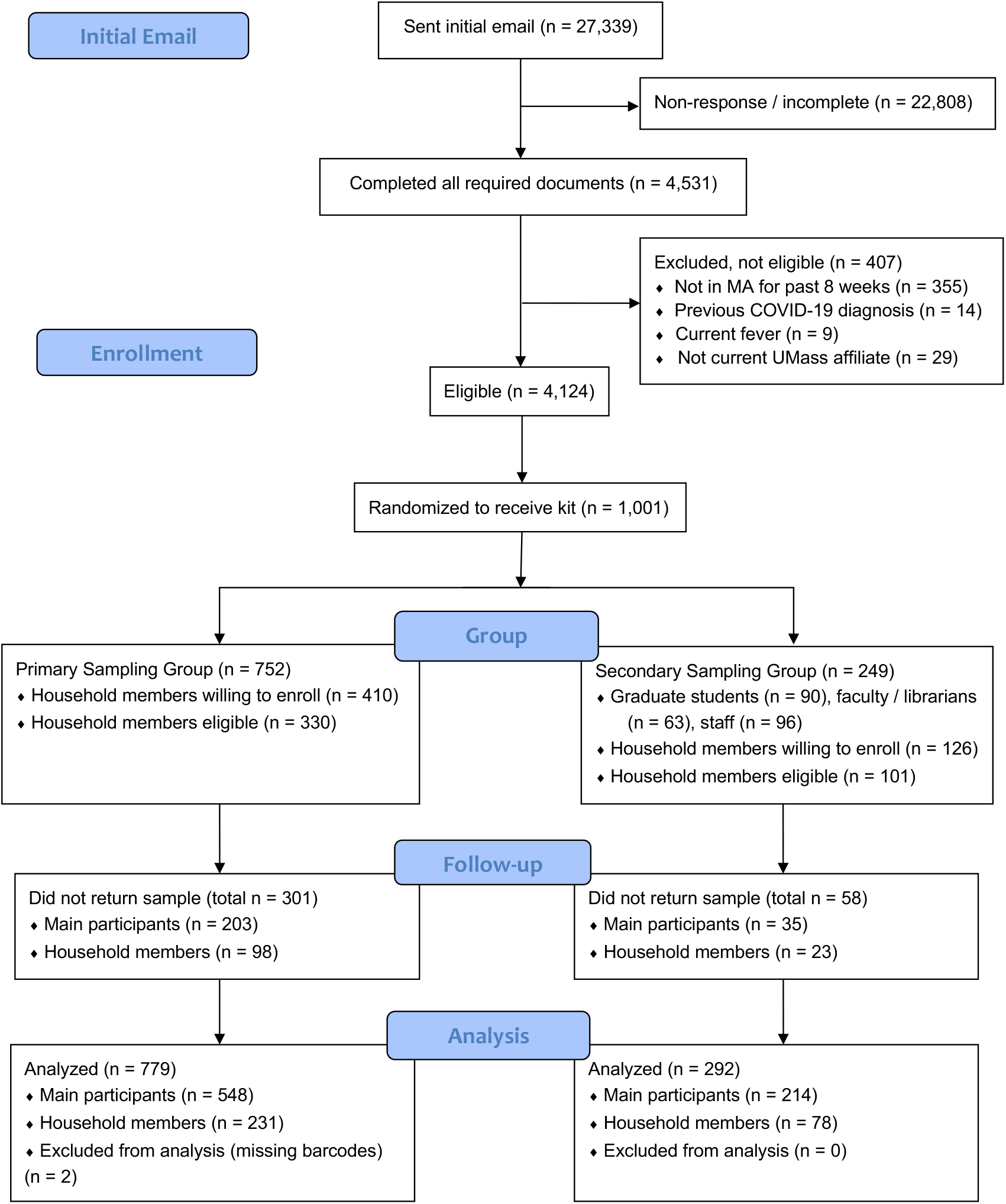
Participant inclusion (CONSORT) enrollment, SARS-CoV-2 serosurvey, Massachusetts, USA, Jul-Aug 2020.

### Sample Preparation and ELISA Analysis

Upon receipt of boxes, the sample cards (Whatman^®^ Protein Saver 903) were heat-treated (30 minutes at 56° C); a single blood spot per card was punched (0.25-inch diameter) and transferred to an ELISA plate. Plates were coated with 1 µg/ml of purified RBD diluted in PBS overnight at 4C and blocked with Tris-Buffered Saline with 0.1% Tween 20 (TBST) containing 5% non-fat dry milk. DBS were eluted in 500 µl of TBST overnight at 4C and 50 μl of each sample was added to the ELISA plate preloaded with 50 µl of TBST containing 2% non-fat dry milk. Samples were then assayed for SARS-CoV-2 antibodies according to published protocols.^18,19^ The RBD protein was produced in-house via transfection of HEK293T cells using polyethylenimine (Plasmid was a generous gift from Pr. F. Kramer mount Sinai School of Medicine). Batches were control for purity by SDS-PAGE followed by Coomassie staining and ELISA using an anti His-tag monoclonal antibody. Optical densities were read at 405 nm, and each 96-well plate contained seven negative controls and one positive control (serum from PCR-confirmed case at 1/100 dilution). Samples were tested against IgG and positive samples were confirmed and tested with anti IgM antibodies. Optical density values were normalized to the mean optical density of negative controls daily.

### Data Analysis

#### Sample size and power

The study was designed to assess seropositivity within the primary sampling group with sufficient precision to inform policy. With 750 persons, and an assumed 5% positivity, the 95% CI for this estimate is 3.6% to 6.9%. Within the five emergency response subregions, at 5% seropositivity, the survey is powered for a precision of 2.3% to 10.2%. The secondary sampling group (n= 250) sample size was based on logistic limitations, but was powered to a precision of 2.8% to 8.8%. All confidence intervals are binomial exact, without adjustments for study design effects or non-response.

#### Analysis of serology data

Finite mixture models were used to determine seropositivity cutoffs. These latent-class models estimate “breakpoints” for seropositive and seronegative subpopulations, and have been applied to a range of pathogen serosurvey data, including rubella, pertussis, and parvovirus.^20–22^ From this analysis, all samples with IgG optical density ratio ≥ 2.49-fold above daily background were considered positive for SARS-CoV-2.

#### Adjusted estimates

All reported prevalences and prevalence ratio estimates are adjusted with non-response weights, which were estimated using inverse weighting. Briefly, logistic regression models were used to calculate propensity scores for each individual in the sample using reported gender and race categories. These were transformed to probabilities; a small number of individuals had extremely large weights due to sparse strata; these weights were truncated at 1/0.02.^23^ Weights were then used for all prevalence and prevalence ratio estimations using the *survey* package in R.^24^ The primary sampling group sample was self-weighting due to probability-proportional to population size sampling. Sampling weights were not used in the secondary sampling group as selection used simple random sampling.

#### Multivariable analyses for prevalence ratios

Prevalence ratios were estimated to assess factors associated with seropositivity, with separate Poisson models^25^ for both of the two sampling groups, with robust (sandwich) errors to address clustering within households.

Bivariate analyses were performed for each factor separately. All variables with a p-value < 0.20 based on bivariate association with outcome were further evaluated for inclusion in final models. All final models were adjusted for age (continuous), race, and gender (see Table 1). Due to several very sparse categories, some were combined in final models. Specifically, all race/ethnicity categories and all geographic regions were not included in analysis of the secondary sampling group due to unstable estimates.

**Table 1.** Demographics of study populations, SARS-CoV-2 serology surveys in university-affiliated populations, Massachusetts Jul-Aug, 2020.

| Characteristic | Primary Sampling Group<br>(N=779) | Secondary Sampling Group<br>(N=292) |
| --- | --- | --- |
| Gender |  |  |
| Female | 499 (64.1%) | 154 (52.7%) |
| Male | 274 (35.2%) | 136 (46.6%) |
| Gender Diverse | 5 (0.6%) | 2 (0.7%) |
| Missing | 1 (0.1%) | 0 (0.0%) |
| Race |  |  |
| AIAN | 17 (2.2%) | 1 (0.3%) |
| Asian | 78 (10.0%) | 34 (11.6%) |
| Black | 12 (1.5%) | 3 (1.0%) |
| Hispanic | 36 (4.6%) | 9 (3.1%) |
| Multiple | 37 (4.8%) | 11 (3.8%) |
| White | 545 (70.0%) | 217 (74.3%) |
| Missing | 54 (6.9%) | 17 (5.8%) |
| Age |  |  |
| Mean | 29.9 | 41.6 |
| Median | 21 | 39 |
| Range | 18 – 75 | 21 – 75 |
| Education |  |  |
| HS/GED | 102 (13.1%) | 5 (1.7%) |
| Some College | 483 (62.0%) | 24 (8.2%) |
| BA/BS | 117 (15.0%) | 78 (26.7%) |
| More Than BA/BS | 74 (9.5%) | 183 (62.7%) |
| Missing | 3 (0.4%) | 2 (0.7%) |
| Essential Worker |  |  |
| No | 533 (68.4%) | 224 (76.7%) |
| Yes | 195 (25.0%) | 51 (17.5%) |
| Missing | 51 (6.6%) | 17 (5.8%) |
| Self-reported attitude<br>about COVID-19 |  |  |
| Strongest fear | 135 (17.3%) | 72 (24.7%) |
| Somewhat fearful | 389 (49.9%) | 122 (41.8%) |
| Neutral/Missing | 139 (17.8%) | 63 (21.6%) |
| Somewhat not fearful | 86 (11.0%) | 23 (7.9%) |
| Not fearful | 30 (3.9%) | 12 (4.1%) |
| Self-reported febrile<br>illness since February | No: 534 (68.6%)<br>Yes: 188 (24.1%)<br>Missing: 57 (7.3%) | No: 224 (76.7%)<br>Yes: 53 (18.2%)<br>Missing: 15 (5.1%) |
| Self-reported care | No: 112 (59.6%) | No: 32 (60.4%) |
| Seeking (if illness since Feb) | Yes: 75 (39.9%)<br>Missing: 1 (0.5%) | Yes: 21 (39.6%)<br>Missing: 0 (0.0%) |

\*Notes: AIAN= American Indian/Alaska Native. The primary sampling group includes UMass undergraduates and household members, and the secondary sampling group includes UMass affiliated faculty, staff, and graduate students and household members.

Model parsimony was evaluated using AIC/BIC and all tests were two-tailed, with α = 0.05. R version 4.0.3 and SAS version 9.4 (SAS Institute, Inc, Cary, NC, USA) were used for analysis.

## Results

A total of 1,001 individuals were enrolled into the study; this included 752 undergraduate students, 90 graduate students, 63 faculty/librarians, and 96 staff members (Figure 2). Seventy-six percent of these (n=762) returned blood samples for analysis; 548 in the primary sampling group, and 214 in the secondary sampling group. Of the 548 participants in the primary sampling group, 230 enrolled a household member. One household member submitted a sample without the sample of the main participant, bringing the total number of undergraduate household members to 231. Of the 214 participants in the secondary sampling group, 78 enrolled a household member. Two returned samples were excluded from analysis due to unlinkable samples. A total of 1,071 samples were included in the final analyses: 762 main participants and 309 household members (Figure 2).

Demographic characteristics of both sampling groups are presented in Table 1. Race categories do not total to 100% due to non-response and multiple possible answers. Age, gender, and essential worker status were similar between those invited to participate and those who completed the study (<u>Supplemental Table 1</u>).

Of the total 1,071 samples tested, 46 were positive for SARS-CoV-2 antibodies. Demographic results are stratified by IgG serostatus (Table 3); no samples showed evidence for IgM positivity. Seropositivity was low-to-moderate across the survey groups, with several important exceptions. Variation is apparent by sex, race, and across geographic regions; however, several strata have wide confidence intervals due to small sample sizes.

Of the 779 primary sampling group participants and their household members, 36 were positive for SARS-CoV-2 antibodies. This corresponds to an overall seroprevalence of 5.3% (95% CI: 3.1 – 7.5) of the population after adjustment for nonresponse and geographic location. In the secondary sampling group, of the 292 graduate students, staff, librarians, faculty members, and their household members, ten (adjusted 4.0 %, 95% CI: 1.6 - 6.5) had evidence for prior SARS-CoV-2 infection. (Table 2). Results were also further stratified by UMass affiliate vs. household member. Of the 548 undergraduate students in the primary sampling group, 27 were positive for SARS-CoV-2 IgG antibodies (population positivity rate of 5.3% (95% CI: 3.1 – 7.6%). Of the 231 household members of undergraduate participants, nine (adjusted 5.2%, 95% CI: 1.2 - 9.2) were positive for SARS-CoV-2 IgG antibodies. In the secondary sampling group, eight University affiliates (adjusted 4.3 %, 95% CI: 1.3 - 7.3) were positive for SARS-CoV-2 IgG antibodies. Of the household members in the secondary group, 2 were seropositive, with a weighted seroprevalence of (3.3 %, 95%CI: 0.0 – 7.8%). (<u>Supplemental Table 2</u>). The overall distributions of measured IgG log normal optical density ratios by subgroups are broadly similar (Supplemental Figure 1).

**Table 2.**
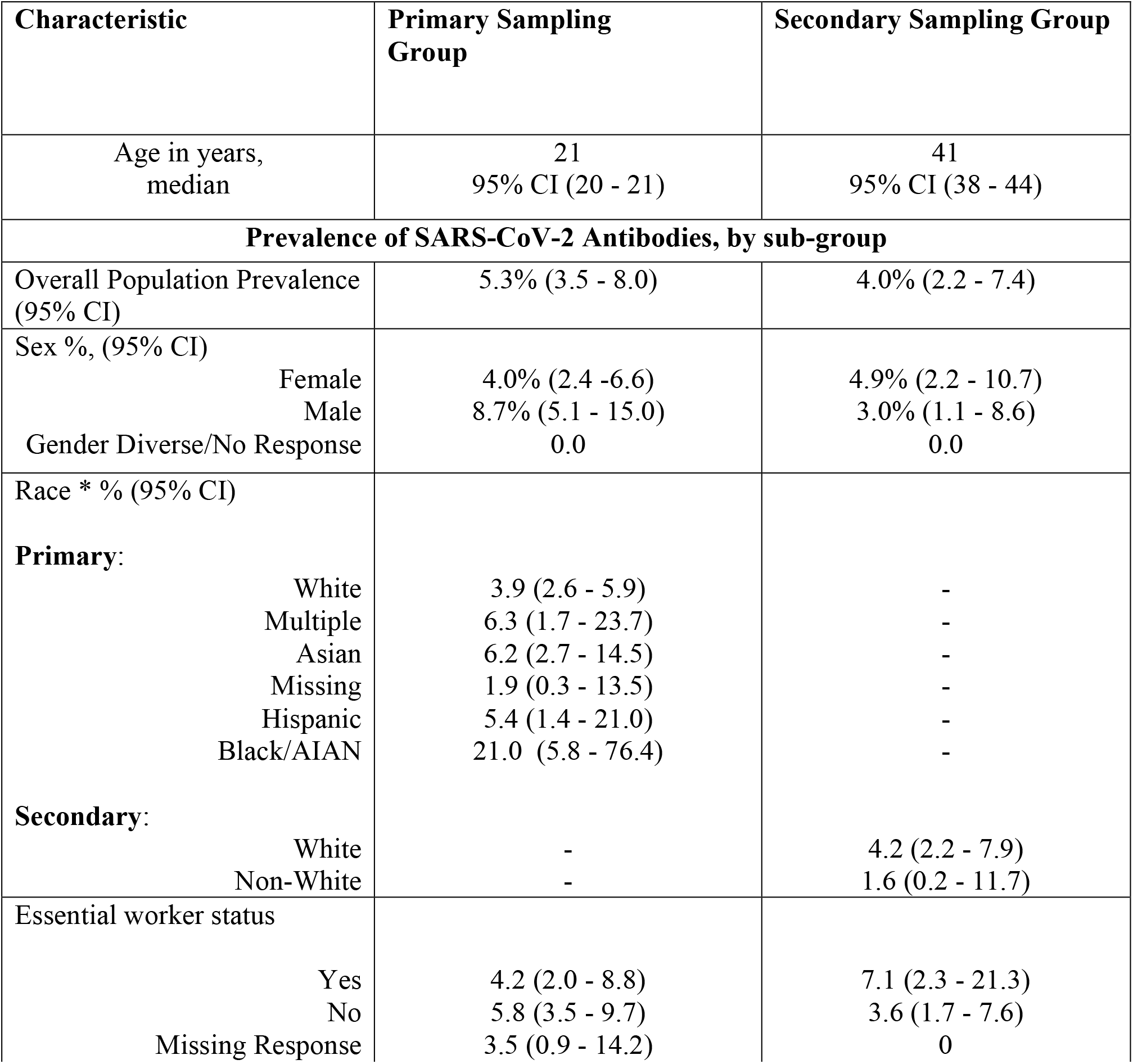
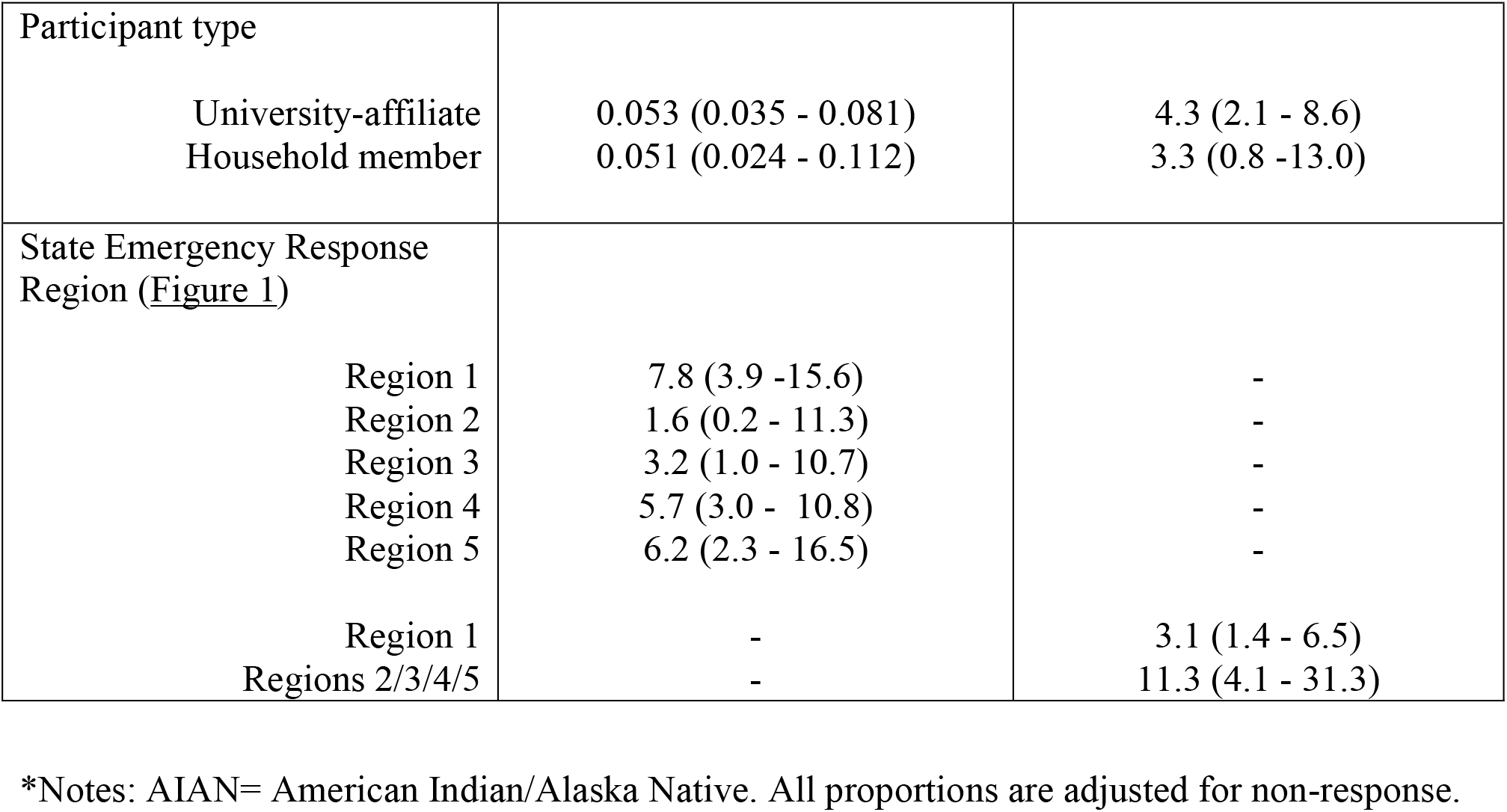
Weighted seropositivity by main demographic variables, MA USA, Jul-Aug 2020.

| Characteristic | Primary Sampling Group | Secondary Sampling Group |
| --- | --- | --- |
| Age in years, median | 21<br>95% CI (20 - 21) | 41<br>95% CI (38 - 44) |
| **Prevalence of SARS-CoV-2 Antibodies, by sub-group** |  |  |
| Overall Population Prevalence (95% CI) | 5.3% (3.5 - 8.0) | 4.0% (2.2 - 7.4) |
| Sex %, (95% CI) |  |  |
| Female | 4.0% (2.4 - 6.6) | 4.9% (2.2 - 10.7) |
| Male | 8.7% (5.1 - 15.0) | 3.0% (1.1 - 8.6) |
| Gender Diverse/No Response | 0.0 | 0.0 |
| Race * % (95% CI) |  |  |
| **Primary:** |  |  |
| White | 3.9 (2.6 - 5.9) | - |
| Multiple | 6.3 (1.7 - 23.7) | - |
| Asian | 6.2 (2.7 - 14.5) | - |
| Missing | 1.9 (0.3 - 13.5) | - |
| Hispanic | 5.4 (1.4 - 21.0) | - |
| Black/AIAN | 21.0 (5.8 - 76.4) | - |
| **Secondary:** |  |  |
| White | - | 4.2 (2.2 - 7.9) |
| Non-White | - | 1.6 (0.2 - 11.7) |
| Essential worker status |  |  |
| Yes | 4.2 (2.0 - 8.8) | 7.1 (2.3 - 21.3) |
| No | 5.8 (3.5 - 9.7) | 3.6 (1.7 - 7.6) |
| Missing Response | 3.5 (0.9 - 14.2) | 0 |
| Participant type |  |  |
| University-affiliate | 0.053 (0.035 - 0.081) | 4.3 (2.1 - 8.6) |
| Household member | 0.051 (0.024 - 0.112) | 3.3 (0.8 -13.0) |
| State Emergency Response<br>Region (Figure 1) |  |  |
| Region 1 | 7.8 (3.9 -15.6) | - |
| Region 2 | 1.6 (0.2 - 11.3) | - |
| Region 3 | 3.2 (1.0 - 10.7) | - |
| Region 4 | 5.7 (3.0 - 10.8) | - |
| Region 5 | 6.2 (2.3 - 16.5) | - |
| Region 1 | - | 3.1 (1.4 - 6.5) |
| Regions 2/3/4/5 | - | 11.3 (4.1 - 31.3) |

\*Notes: AIAN= American Indian/Alaska Native. All proportions are adjusted for non-response.

After adjustments for age, gender, region, and self-reported febrile illness since February 2020, the factor with the strongest association with seropositivity in the primary sampling group was Black or AIAN Race (PR = 4.49, 95% CI = 1.57,12.9) (Table 3). This indicates that individuals who report being Black or American Indian / Alaskan Native have a prevalence 3.49 times higher than White individuals after adjustment. Additionally, after adjustments, females and those who are gender diverse were at a significantly lower risk of prior SARS-CoV-2 infection (PR = 0.5; 95% CI = 0.27, 0.92) compared to males. Those who reported a febrile illness in February were more likely to be seropositive than those who were not sick (PR = 2.42, 95% CI = 1.24, 4.75). No significant associations were found across each of the 5 regions in the primary sampling group, however the prevalence of seropositivity was 48% higher in Region 1 compared to Region 4 (PR = 1.48, 95% CI = 0.62, 3.52).

**Table 3.** Multivariable associations for SARS-CoV-2 seropositivity, Primary Sampling Group, MA USA, Jul-Aug 2020.

| Characteristic | Prevalence Ratio | 95% CI | p-value |
| --- | --- | --- | --- |
| Emergency Response Region |  |  |  |
| Region 1 | 1.48 | 0.62, 3.52 | 0.38 |
| Region 2 | 0.34 | 0.05, 2.45 | 0.28 |
| Region 3 | 0.53 | 0.14, 1.96 | 0.34 |
| Region 4 | Reference |  |  |
| Region 5 | 1.02 | 0.35, 2.98 | 0.97 |
| Age (years) | 1.04 | 0.96, 1.12 | 0.33 |
| Gender |  |  |  |
| Male | Reference |  |  |
| Female, Gender diverse, or No response | 0.50 | 0.27, 0.92 | **0.027** |
| Race |  |  |  |
| White | Reference |  |  |
| Multiple | 1.91 | 0.46, 7.98 | 0.38 |
| Asian | 1.66 | 0.66, 4.16 | 0.28 |
| Missing Race | 0.51 | 0.07, 3.71 | 0.51 |
| Hispanic | 1.76 | 0.44, 7.04 | 0.42 |
| Black or AIAN | 4.49 | 1.57, 12.9 | **0.005** |
| Febrile illness since February |  |  |  |
| No | Reference |  |  |
| Yes | 2.42 | 1.24, 4.75 | **0.010** |
| Missing Response | 0.33 | 0.04, 2.45 | 0.28 |
| Other household member |  |  |  |
| no | Reference |  |  |
| yes | 0.29 | 0.01, 7.69 | 0.46 |

Within the secondary sampling population, after adjustments for age, race, gender, region, household member and self-reported febrile illness since February (Table 4), participants who reported residing in Region 2, 3, 4, or 5 had greater than 4 times higher prevalence of SARS-CoV-2 antibodies as compared to those who resided in Region 1 (PR = 4.08, 95% CI = 1.09, 15.33). No other factors included in the model were significantly associated with seropositivity.

**Table 4.** Multivariable associations for SARS-CoV-2 seropositivity, Secondary Sampling Group, Massachusetts, USA, Jul-Aug 2020.

| Characteristic | Prevalence Ratio | 95% CI | p-value |
| --- | --- | --- | --- |
| Age (years) | 1.03 | 0.99, 1.07 | 0.17 |
| Gender |  |  |  |
| Male | Reference | - | - |
| Female or Gender Diverse | 1.35 | 0.43, 4.31 | 0.61 |
| Race |  |  |  |
| White | Reference | - | - |
| All Other/Multiple/Missing | 0.58 | 0.07, 4.86 | 0.62 |
| Febrile illness since February |  |  |  |
| No | Reference | - | - |
| Yes | 2.56 | 0.68, 9.67 | 0.17 |
| Missing Response | 2.35 | 0.21, 26.73 | 0.49 |
| Emergency Response Region |  |  |  |
| Region 1 | Reference | - | - |
| Regions 2/3/4/5 | 4.08 | 1.09, 15.33 | **0.039** |
| Other household member |  |  |  |
| No | Reference | - | - |
| Yes | 0.70 | 0.18, 2.72 | 0.61 |

## Discussion

This mail-based serosurvey of two university-affiliated populations across Massachusetts in July and August 2020 found an estimated seroprevalence of ∼5% of antibodies to SARS-CoV-2. These results indicate that even with extensive morbidity and mortality across the state at the time of sampling, there had been limited exposure to SARS-CoV-2 at a population-level. This estimated seroprevalence is lower than that detected with concurrent community-based studies in other states. An estimated 14.3% of the United States population had been previously infected with SARS-CoV-2 by November 2020, as estimated in a pooled analysis of multiple seroprevalence surveys.^26^

Our estimates are substantially lower than some models of COVID-19 seroprevalence in Massachusetts. One model estimates a seroprevalence of 16.2% (no CIs provided) on Jul 27, 2020 (closest modeled date). These estimates are nearly double our measured seroprevalence with inclusion of 110,000 confirmed cases at that date (ca. 1.5%)^27^. These differences might be caused by a number of reasons, including a non-representative population by age or geographic range, or waning of antibody titers. Without CIs, we are unable to evaluate coverage outside the reported point estimate. However, alternate nowcasting estimates suggest a total statewide attack rate on July 31, 2020 of 6.9% (95% CrI: 5.5 – 8.4%) in Massachusetts^28^. Our results are closely aligned with these estimates.

Within the surveyed groups, approximately 24% of the primary sampling group and 18% of the secondary sampling group reported illness since February. Febrile illness since February was associated with an increased prevalence of seropositivity in both sampling groups, but after multivariable analysis this association was found in only the primary sampling group. These results reinforce results from other studies: asymptomatic illness is an important contributor to observed force of infection; and important limitations of testing availability at the time of survey.

Differing antibody dynamics have been reported in other studies. A number of studies have found sustained antibody levels for over 3 months,^29,30^ while others have found IgG levels can remain 6 months or more.^31–33^ An additional study has reported rapid waning of routine serological markers in individuals who had lower initial antibody responses.^34^. Only 7.1% of those with high titers at baseline seroreverted to a level below the threshold for positivity within 60 days, compared to 64.9% of those with lower titers at baseline.^34^ Evidence for IgM seropositivity was not detected in any of IgG positive samples, which is consistent with results from other surveys studies that included asymptomatic or subclinical populations due to rapidly waning titers.^32,35^ Studies have shown that IgM levels decline more rapidly after infection than IgA and IgG levels, ^30,36,37^, and this is especially apparent with asymptomatic and sub-clinical infections.^32,35^

Trends in the patterns of SARS-CoV-2 antibody levels vary greatly depending on the timing of sampling and severity of disease.^31,32,35^ Seroconversion times vary depending on the study, but one study found a median time-to-seroconversion for IgM of 8 days and median seroconversion for IgG of 10 days. Additionally, the SARS-CoV-2 IgG response generally begin around 10-15 days after symptom onset.^2^ For this reason, repeated serial sampling, repeated serial sampling of convalescent populations should be prioritized to more fully understand the dynamics of immune response.

SARS-CoV-2 seropositivity was associated with minority race status in this survey. While the total number of non-white participants was limited, the large effect size reinforces other studies suggesting that marginalized communities have been and continue to be excessively impacted by the pandemic. Results from the primary sampling group analysis suggest that self-reported Black race is a risk factor for previous SARS-CoV-2 infection, which is consistent with findings from other studies.^38–40^ A number of factors may play a role in this significant association including the fact that Black individuals are more likely to work in frontline industries or live in areas with a higher population density.^41^ No parallel associations were found in the analysis of the secondary sampling group due to limited sample size in some strata. In the secondary sampling group, the aggregation of Race categories into White race and Non-white race likely obscured meaningful associations between Race and exposure.

Results from the primary sampling group showed higher prevalence of IgG seropositivity among males. After adjusting for age, race, and region, male gender was a significant risk factor for evidence of SARS-CoV-2 infection. Other studies have similarly found that males have higher rates of infection than females for asymptomatic infections^42,43^ These findings may reflect differences in care seeking behavior (biased recruitment), true biological differences, or differences in health behaviors such as smoking, alcohol use, and COVID-19 prevention measures.^44^ This association was not observed in the secondary sampling group.

The primary sampling group also showed increased risk of seropositivity with self-reported illness since February 2020; this association was not observed in analyses of the secondary sampling group. This finding may indicate that some of the participants in our study were not strictly asymptomatic and were simply unable to obtain a COVID-19 test due to limited availability during the beginning of the pandemic.

This study was population-based and had broad eligibility criteria but is subject to several limitations. The exclusion of persons with confirmed diagnoses and any current symptoms (due to biosafety concerns) also inherently limited capture of sub-clinical infections. As such, the estimates are likely a lower bound. However, participants who suspected they may have been previously infected with SARS-CoV-2 might be more likely to participate compared to those that were not concerned with prior infection. This is a pervasive issue in community-based studies, where characteristics of those who volunteer to participate in community-based research differ from the general population.^45^

Randomization after a three-week enrollment period helped to address this limitation, as using only the first participants to volunteer could have biased the sample to include those who were most motivated to receive their antibody test results. If participants were more motivated to receive their results because they thought they suspected a prior exposure to SARS-CoV2, this would have inflated the observed prevalence of seropositivity in the study population.

Another limitation of the study is the self-reported response of the lack of a prior COVID-19 diagnosis and current fever. It is possible that some participants shielded their answer and submitted samples for analysis without meeting the eligibility criteria; this would have inflated our estimation of seroprevalence in asymptomatic groups, Thirdly, the limited number of non-White, and gender-diverse participants also limited some analyses. Fourthly, while multiple studies have validated DBS sampling for SARS-CoV-2,^46^ waning antibodies in asymptomatic individuals could be below the limited of detection of the ELISA assay. Finally, generalizability is limited due to the recruitment of a university-affiliated population in a relatively restricted geographic area.^8^

This serosurvey estimates prevalence of prior SARS-CoV-2 infections in a university-affiliated population in Massachusetts. Risk factors for IgG seropositivity included recent self-reported febrile illness, minority race status, and male gender. This study provides estimates of seroprevalence in Massachusetts after a ‘first wave’ of SARS-CoV-2 infections in the spring of 2020. Repeat seroprevalence studies in this population could provide estimates in changes of seropositivity rates given subsequent waves of SARS-CoV-2 transmission. This study reinforces the critical need for targeted serosurveys in highest-risk and marginalized communities, both in Massachusetts, and nationwide.

## Data Availability

All data will be available at osf.io.

## Acknowledgements

We would like to express our appreciation to all of the individuals who have enabled the completion of this study. Rob Leveille and Charlie Apicella of the UMass Mail and Distribution Services have facilitated the label printing, outgoing shipments, and incoming shipments for over 1,000 sample boxes. Sujitha Chandra Kumar, Vincent Lee, and Pratik Patel have been valuable members of the REDCap support team. Kimberly Tremblay, Jesse Mager, Katherine Dorfman, Pa Tamba Ngom, and Ryan Kurtz have graciously allowed us to utilize laboratory space for box assembly and sample processing. These individuals have worked tirelessly to support this study despite massive pandemic-related logistical challenges and tight deadlines. We are very grateful for their support.

