## Supplemental data- Table S1 and Figure S1 for "Serological surveys to estimate cumulative incidence of SARS-CoV-2 infection in adults (Sero-MAss study), Massachusetts, July-August 2020: a mail-based cross-sectional study"

### Supplemental Information

Supplemental Table 1: Comparison of characteristics of participants selected for randomization and those not selected for randomization, SARS-CoV-2 serosurvey, Massachusetts, USA, Jul-Aug 2020.

| | Primary Sampling Group (undergraduates) | | Secondary sampling group (employees) | |
|---|---|---|---|---|
| | Not Randomized | Randomized | Not Randomized | Randomized |
| n | 869 | 752 | 2253 | 249 |
| Age (mean (SD)) | 20.64 (3.09) | 20.18 (1.79) | 42.05 (13.53) | 40.69 (13.88) |
| Gender (%) | | | | |
| Unknown | 0 (0.0) | 0 (0.0) | 1 (0.0) | 0 (0.0) |
| Female | 525 (60.4) | 453 (60.2) | 1262 (56.0) | 147 (59.0) |
| Gender diverse | 12 (1.4) | 8 (1.1) | 49 (2.2) | 3 (1.2) |
| Male | 332 (38.2) | 291 (38.7) | 941 (41.8) | 99 (39.8) |
| Febrile Illness during February (%) | | | | |
| Unknown | 1 (0.1) | 2 (0.3) | 6 (0.3) | 0 (0.0) |
| No | 568 (65.4) | 499 (66.4) | 1658 (73.6) | 189 (75.9) |
| Not sure | 75 (8.6) | 57 (7.6) | 154 (6.8) | 16 (6.4) |
| Yes | 225 (25.9) | 194 (25.8) | 435 (19.3) | 44 (17.7) |
| Education (%) | | | | |
| Unknown | 4 (0.5) | 3 (0.4) | 6 (0.3) | 1 (0.4) |
| BA/BS | 15 (1.7) | 14 (1.9) | 509 (22.6) | 64 (25.7) |
| High school / GED | 90 (10.4) | 108 (14.4) | 46 (2.0) | 2 (0.8) |
| More than BA/BS | 2 (0.2) | 0 (0.0) | 1528 (67.8) | 166 (66.7) |
| Prefer not to answer | 1 (0.1) | 0 (0.0) | 13 (0.6) | 1 (0.4) |
| Some college | 757 (87.1) | 627 (83.4) | 151 (6.7) | 15 (6.0) |
| Response to, "I am afraid of COVID-19" (%) | | | | |
| Unknown | 1 (0.1) | 2 (0.3) | 2 (0.1) | 0 (0.0) |
| Neither agree nor disagree | 199 (22.9) | 141 (18.8) | 399 (17.7) | 51 (20.5) |
| Somewhat agree | 390 (44.9) | 374 (49.7) | 1051 (46.6) | 100 (40.2) |
| Somewhat disagree | 115 (13.2) | 99 (13.2) | 153 (6.8) | 22 (8.8) |
| Strongly agree | 121 (13.9) | 104 (13.8) | 573 (25.4) | 66 (26.5) |
| Strongly disagree | 43 (4.9) | 32 (4.3) | 75 (3.3) | 10 (4.0) |

S-Fig 1. Distributions of IgG log normal optical densities (OD) ratios, by subgroups, SARS-CoV-2, Massachusetts, USA, Jul-Aug 2020.

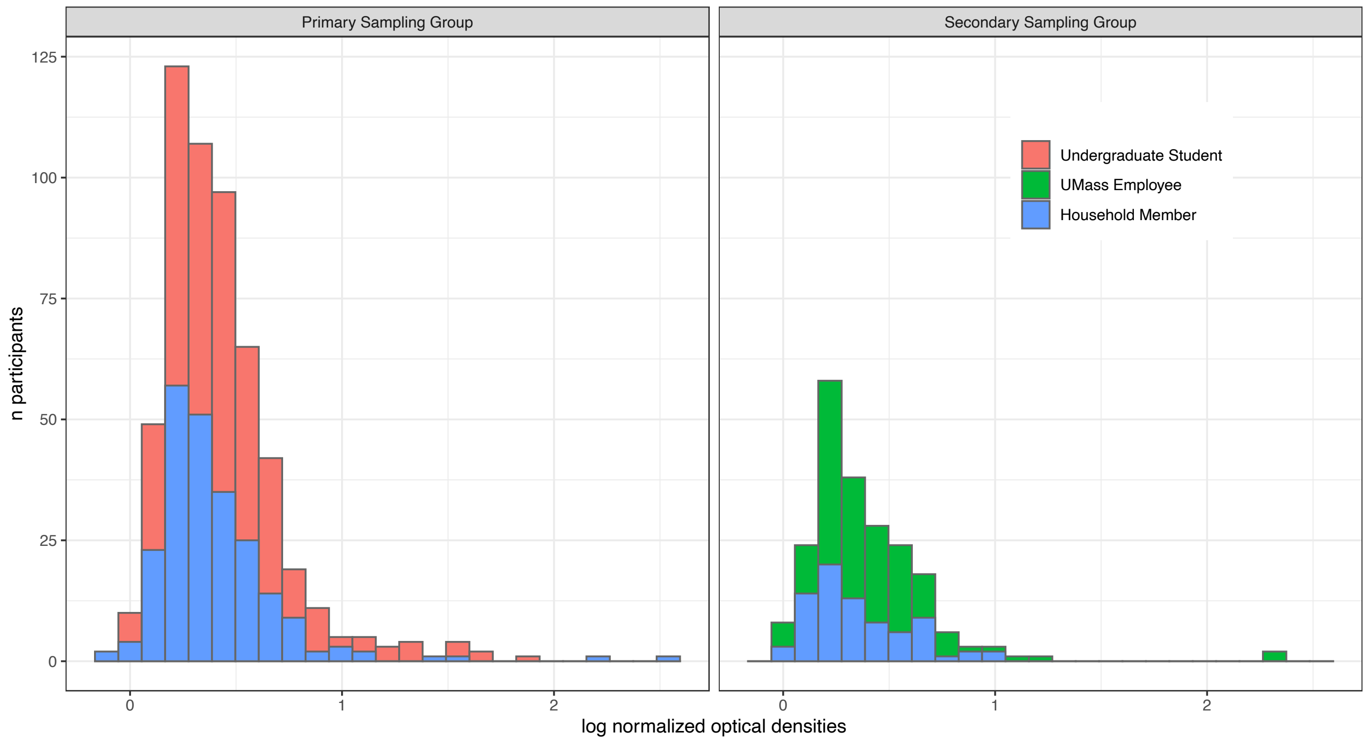
